# Children develop robust and sustained cross-reactive spike-specific immune responses following SARS-CoV-2 infection

**DOI:** 10.1101/2021.04.12.21255275

**Authors:** Alexander C. Dowell, Megan S. Butler, Elizabeth Jinks, Gokhan Tut, Tara Lancaster, Panagiota Sylla, Jusnara Begum, Rachel Bruton, Hayden Pearce, Kriti Verma, Nicola Logan, Grace Tyson, Eliska Spalkova, Sandra Margielewska-Davies, Graham S. Taylor, Eleni Syrimi, Frances Baawuah, Joanne Beckmann, Ifeanyichukwu Okike, Shazaad Ahmad, Joanna Garstang, Andrew J Brent, Bernadette Brent, Georgina Ireland, Felicity Aiano, Zahin Amin-Chowdhury, Samuel Jones, Ray Borrow, Ezra Linley, John Wright, Rafaq Azad, Dagmar Waiblinger, Chris Davis, Emma Thomson, Massimo Palmarini, Brian J. Willett, Wendy S. Barclay, John Poh, Vanessa Saliba, Gayatri Amirthalingam, Kevin E Brown, Mary E Ramsay, Jianmin Zuo, Paul Moss, Shamez Ladhani

**Affiliations:** Institute of Immunology & Immunotherapy, College of Medical and Dental Sciences, University of Birmingham, Birmingham, B15 2TT, UK; MRC-University of Glasgow Centre for Virus Research, 464 Bearsden Road, Glasgow G61-1QH, UK; Public Health England, 61 Colindale Avenue, London NW9 5EQ, UK; East London NHS Foundation Trust, 9 Allie Street, London E1 8DE, UK; University Hospitals of Derby and Burton NHS Foundation Trust, Uttoxeter New Road, Derby DE22 3NE, UK; Manchester University NHS Foundation Trust, Oxford Road, Manchester M13 9WL, UK; Birmingham Community Healthcare NHS Trust, Holt Street, Aston B7 4BN, UK; Oxford University Hospitals NHS Foundation Trust, Old Road, Oxford OX3 7HE; University of Oxford, Wellington Square, Oxford OX1 2JD, UK; Public Health England, Manchester Royal Infirmary, Manchester, United Kingdom; Bradford Institute for Health Research, Bradford Teaching Hospitals NHS Foundation Trust, Bradford, BD9 6RJ, UK; Department of Infectious Disease, Imperial College, London, UK; Paediatric Infectious Diseases Research Group, St. George’s University of London, London, UK

## Abstract

SARS-CoV-2 infection is generally mild or asymptomatic in children but the biological basis for this is unclear. We studied the profile of antibody and cellular immunity in children aged 3-11 years in comparison with adults. Antibody responses against spike and receptor binding domain (RBD) were high in children and seroconversion boosted antibody responses against seasonal Beta-coronaviruses through cross-recognition of the S2 domain. Seroneutralisation assays against alpha, beta and delta SARS-CoV-2 variants demonstrated comparable neutralising activity between children and adults. T cell responses against spike were >2-fold higher in children compared to adults and displayed a T_H_1 cytokine profile. SARS-CoV-2 spike-specific T cells were also detected in many seronegative children, revealing pre-existing responses that were cross-reactive with seasonal Alpha and Beta-coronaviruses. Importantly, all children retained high antibody titres and cellular responses at 6 months after infection whilst relative antibody waning was seen in adults. Spike-specific responses in children also remained broadly stable beyond 12 months. Children thus distinctly generate robust, cross-reactive and sustained immune responses after SARS-CoV-2 infection with focussed specificity against spike protein. These observations demonstrate novel features of SARS-CoV-2-specific immune responses in children and may provide insight into their relative clinical protection. Furthermore, this information will help to guide the introduction of vaccination regimens in the paediatric population.

## Introduction

The SARS-CoV-2 pandemic has resulted in over 4.2 million deaths to date and the most significant determinant of outcome is age at the time of primary infection^1^. SARS-CoV-2 infection in children is generally asymptomatic or mild and contrasts with high rates of hospitalisations and deaths among older adults ^2^. As such, there is interest in understanding the profile of the immune response to SARS-CoV-2 in children. Such studies are limited to date but have reported reduced magnitude of both antibody and cellular responses in comparison to adults, and an absence of nucleocapsid-specific antibody responses during or early post-infection ^3, 4, 5, 6^. One unique feature of SARS-CoV-2 infection in children is the development of a rare complication known as pediatric inflammatory multisystem syndrome temporally associated with SARS-CoV-2 (PIMS-TS), also known as paediatric multisystem inflammatory syndrome (MIS-C), which shares features with Kawasaki disease and toxic shock syndrome ^7, 8^. MIS-C develops approximately 2-4 weeks after infection in children with a median age of 9 years ^9^. The immunological basis for this condition remains unclear but is characterised by diffuse endothelial involvement and broad autoantibody production ^10^.

One potential determinant of differential immune responses to SARS-CoV-2 across the life course may be the timing of exposure to the four additional endemic human coronaviruses (HCoV). These comprise the Beta-coronaviruses OC43 and HKU-1, which have 38% and 35% amino acid homology with SARS-CoV-2, as well as the more distantly related Alpha-coronaviruses NL63 and 229E, each with around 31% homology ^11^. These coronaviruses cause frequent mild childhood infections and antibody seroconversion occurs typically before the age of 5 years. Infection with one of the Alpha or Beta-coronaviruses provides short-term immunity against re-infection from coronaviruses and is believed to represent transient cross-reactive immunity within the subtypes ^12, 13, 14^. As such, recent HCoV infection might pre-sensitize children against SARS-CoV-2 infection and may explain cross-reactive SARS-CoV-2-neutralising antibodies in some seronegative children ^15^. Immune responses against HCoV are retained throughout life but do not provide sterilising immunity ^13^. Consequently, recurrent infections are common, generating concern that a similar pattern will be observed after SARS-CoV-2 infection.

COVID-19 vaccines are now being administered widely to adult populations and are also being delivered to children in some countries. It is therefore imperative to understand the profile of SARS-CoV-2-specific immune responses in children after natural infection in order to inform vaccination strategy. Here we provide a comprehensive characterisation of the convalescent humoral and cellular immune response in a cohort of 91 primary school-aged children compared with 154 adults taking part in the SARS-CoV-2 Surveillance in School Kids (sKIDs) study ^16^. We demonstrate a markedly different profile of immune response after SARS-CoV-2 infection in children compared to adults. These findings have potential implications for understanding protective or pathological immune responses to infection in children and may help to guide and interpret COVID-19 vaccination regimens for children.

## Methods

### Sample collection

Public Health England (PHE) initiated prospective SARS-CoV-2 surveillance in primary schools across the UK after they reopened following the easing of national lockdown in June 2020. The protocol for the COVID-19 Surveillance in School KIDs (sKIDs) is available online (https://www.gov.uk/guidance/covid-19-paediatric-surveillance) ^16^. Surveillance comprised two arms, one involving weekly swabbing of primary school students and staff for SARS-CoV-2 infection (from June to mid-July 2020) and the other comprising swabbing and blood sampling taken in 3 rounds: beginning (1-19 June) and end (3-23 July) of the second half of the summer term when primary schools were partially re-opened, and after full reopening of all schools in September 2020, at the end of the autumn term (23 November-18 December). Samples for extended humoral and cellular analysis were taken in round 3. Additional samples from children found to be seropositive in round 1 were taken from 21 June-24 July 2021.

For each known SARS-CoV-2 seropositive individual, an age-matched (nearest age in years for students, nearest 10 years for teachers) and sex-matched participant also underwent blood sampling. In total 154 adults and 91 children had sufficient blood sample for serology and cellular responses (Table 1). Convalescent plasma samples were also available from 35 children aged 10-13 years with PCR-confirmed SARS-CoV-2 infection, taken a median of 6 months (range 2-12 months) following PCR result, from the Born in Bradford study ^17^.

**Table 1.**
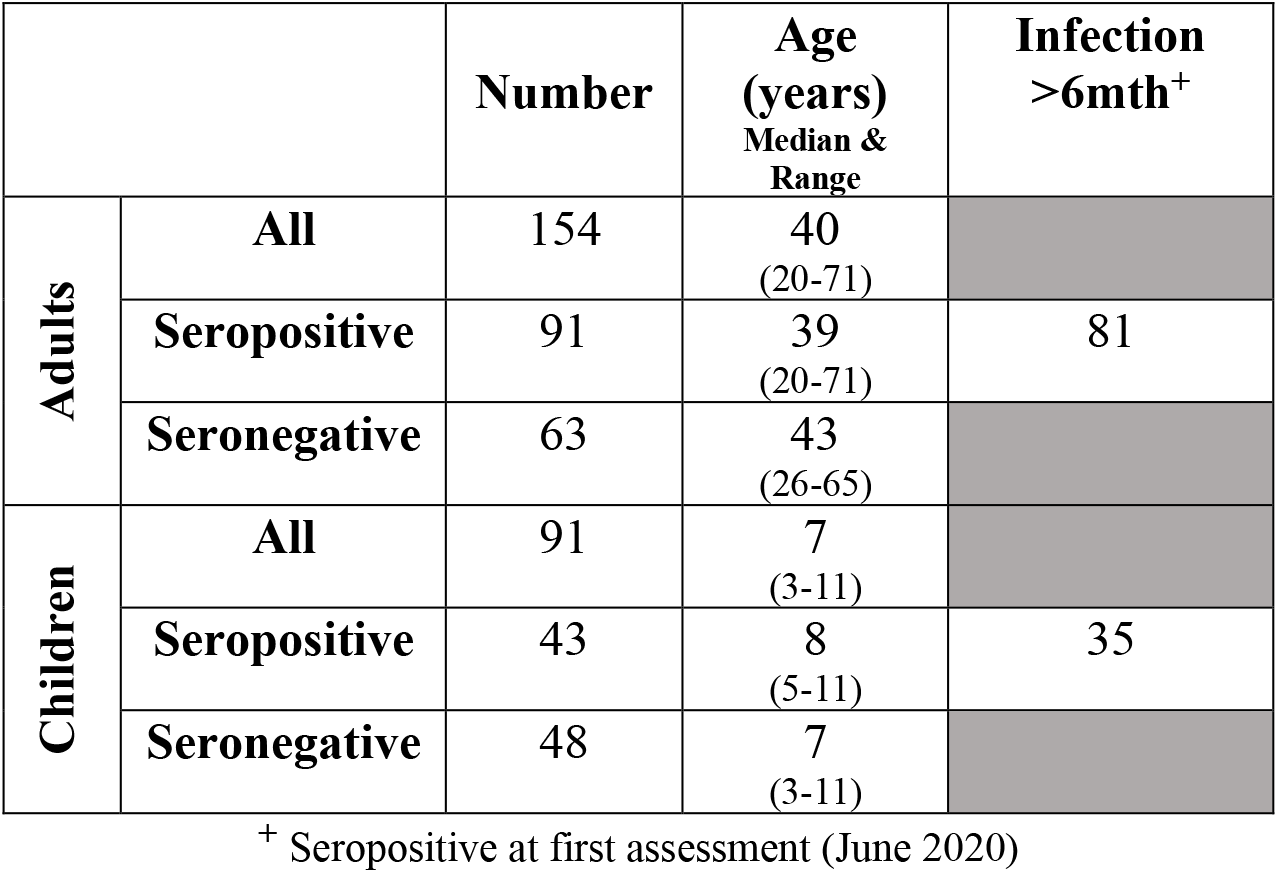
Demographics and serostatus of study participants.

|  |  | Number | Age<br>(years)<br>Median &<br>Range | Infection<br>>6mth<sup>+</sup> |
| --- | --- | --- | --- | --- |
| Adults | All | 154 | 40<br>(20-71) |  |
|  | Seropositive | 91 | 39<br>(20-71) | 81 |
|  | Seronegative | 63 | 43<br>(26-65) |  |
| Children | All | 91 | 7<br>(3-11) |  |
|  | Seropositive | 43 | 8<br>(5-11) | 35 |
|  | Seronegative | 48 | 7<br>(3-11) |  |

<sup>+</sup> Seropositive at first assessment (June 2020)

Pre-pandemic plasma and PBMC were obtained from healthy children as part of an ethically approved study (TrICICL); South of Birmingham Research Ethics Committee (REC: 17/WM/0453, IRAS: 233593).

### PBMC and Plasma Preparation

Blood tubes were spun at 300g for 10mins prior to removal of plasma which was then spun again at 800g for 10mins and stored at -80°C. Remaining blood was diluted 1:1 with RPMI and PBMC isolated on a Sepmate (Stemcell) density centrifugation tube, washed with RPMI and rested in RPMI+10% FBS overnight at 37°C.

### MSD Serology assay

Quantitative IgG antibody titres were measured against trimeric spike (S) protein, nucleocapsid (N) and other coronavirus using MSD V-PLEX COVID-19 Coronavirus Panel 2 (N05368-A1), Coronavirus Panel 7 (N05428A-1) responses to other respiratory viruses were measured using MSD V-PLEX COVID-19 Respiratory Panel 1 (N05358-A1). Multiplex MSD Assays were performed according to manufacturer instructions. Briefly 96-well plates were blocked. Following washing, samples diluted 1:5000 in diluent, as well as reference standard and internal controls, were added to the wells. After incubation plates were washed and detection antibody added. Plates were washed and were immediately read using a MESO TM QuickPlex SQ 120 system. Data were generated by Methodological Mind software and analysed with MSD Discovery Workbench (v4.0) software. Assay Cut-offs in respect of pre-pandemic plasma samples from healthy donors are shown in (Extended Data 1). Cut-offs used are spike; 350 AU/ml, RBD and Nucleocapsid; 600 AU/ml, NTD; 15 AU/ml.

### Total IgG/A/M anti-spike SARS-CoV-2 ELISA

A total GAM anti-SARS-CoV-2 spike ELISA kit ^18^ was purchased from Binding Site (Birmingham, UK). ELISA was performed following the manufacturer’s instructions. Optical density (OD) was compared to a known calibrator and expressed as a ratio to the calibrator. Samples with a ratio above 1.0 were considered seropositive.

### Cross-reactive antibody blocking

Plasma samples were pre-diluted 1:10 with PBS then pre-absorbed by adding an equal volume of either recombinant spike S1 domain (10569-CV-100) or spike S2 (10594-CV-100) (R&D systems) at a concentration of 500ug/ml in PBS, or PBS alone (mock). Samples were incubated at 37°C for 30min. Samples were then diluted to a final dilution of 1:5000 in MSD diluent and run on a MSD V-PLEX COVID-19 Respiratory Panel 2 plate in duplicate.

### RBD-ACE-2 competitive binding assay

The concentration of antibodies which inhibited interaction between RBD and ACE-2 was determined using a SARS-CoV-2 neutralisation assay (Biolegend) following the manufacturer’s instructions. Briefly, plasma or positive control antibody were pre-incubated with biotinylated-Fc-chimera-S1-RBD protein prior to addition of bead bound ACE-2. Binding of RBD to ACE2 was then measured by the addition of streptavidin-PE. Samples were run on a BD LSR-II flow cytometer and analysed using LEGENDplex v8.0 software (Biolegend). Results were related to a known RBD neutralising antibody standard and displayed as ng/ml.

### Live virus and pseudotype-based neutralisation assays

Clinical isolates used in the study were provided by Public Health England and Imperial College London.

A549-ACE2-TMPRSS2 cells ^19^ were seeded at a cell density of 1×10^4^/well in 96-well plates 24hrs before inoculation. Serum was titrated starting at a 1:100 dilution. The specified virus was then incubated at an MOI 0.01 with the Serum for 1hr prior to infection. All wells were performed in triplicate. 72hrs later infection plates were fixed with 8% formaldehyde and stained with Coomassie blue for 30 mins. Plates were washed and dried overnight before quantification using a Celigo Imaging Cytometer (Nexcelom) to measure the staining intensity. Percentage cell survival was determined by comparing the intensity of the staining to uninfected wells.

HEK293, HEK293T, and 293-ACE2 cells were maintained in Dulbecco’s modified Eagle’s medium (DMEM) supplemented with 10% foetal bovine serum, 200mM L-glutamine, 100µg/ml streptomycin and 100 IU/ml penicillin. HEK293T cells were transfected with the appropriate SARS-CoV-2 S gene expression vector in conjunction with lentiviral vectors p8.91 and pCSFLW using polyethylenimine (PEI, Polysciences, Warrington, USA). HIV (SARS-CoV-2) pseudotype-containing supernatants were harvested 48 hours post-transfection, aliquoted and frozen at -80°C prior to use. The SARS-CoV-2 spike glycoprotein expression constructs for Wuhan-Hu-1, B.1.351 (South Africa), B.1.617.2 have been described ^20^. Constructs bore the following mutations relative to the Wuhan-Hu-1 sequence (GenBank: MN908947): B.1.351 – D80A, D215G, L241-243del, K417N, E484K, N501Y, D614G, A701V; B.1.617.2 – T19R, G142D, E156del, F157del, R158G, L452R, T478K, D614G, P681R, D950N. 293-ACE2 target cells were maintained in complete DMEM supplemented with 2µg/ml puromycin.

Neutralising activity in each sample was measured by a serial dilution approach. Each sample was serially diluted in triplicate from 1:50 to 1:36450 in complete DMEM prior to incubation with approximately 1×10^6^ CPS per well of HIV (SARS-CoV-2) pseudotypes, incubated for 1 hour, and plated onto 239-ACE2 target cells. After 48-72 hours, luciferase activity was quantified by the addition of Steadylite Plus chemiluminescence substrate and analysis on a Perkin Elmer EnSight multimode plate reader (Perkin Elmer, Beaconsfield, UK). Antibody titre was then estimated by interpolating the point at which infectivity had been reduced to 90% of the value for the no serum control samples.

### IFN-γ ELISpot

Pepmixes pool containing 15-mer peptides overlapping by 10aa from either SARS-CoV-2 spike S1 or S2 domains and a combined pool of Nucleoprotein (N) and Membrane (M) and Envelope (E) were purchased from Alta Biosciences (University of Birmingham, UK). Overlapping pepmix from Influenza A Matrix Protein 1; California/08/2009(H1N1) ID:C3W5Z8 and Aichi/2/1968 H3N2 ID:Q67157, were purchased from JPT technologies, and combined as a relevant control.

T cell responses were determined using a IFN-γ ELISpot Pro kit (Mabtech) as previously described ^21^. Briefly, fresh PBMC were rested overnight prior to assay and 0.25-0.3×10^6^ PBMC were added in duplicate per well containing either pep-mix, anti-CD3 (positive) or DMSO (negative) control. Samples were incubated for 16-18hrs. Supernatant was harvested and stored at -80°C. Plates were developed following the manufacturer’s instructions and read using an AID plate reader (AID). Cut off values were previously determined ^21^.

### Cross-reactive T cell assay

1.3 ×10^6^ PBMC were peptide-pulsed with SARS-CoV-2 S2 pepmix pool (JPT) at a concentration of 1ug/ml per peptide or an equal volume of DMSO. Cells were then plated into a 48-well plate and cultured in RPMI+10%FBS+Pen/Strep with the addition of 20U/ml IL-2 for 9 days, with frequent media changes. IL-2 was removed 24hrs prior to assay. Cells were washed and divided across four wells of an ELISpot plate and then re-stimulated with either SARS-CoV-2 S2 pepmix or S2 pepmixes from either the Beta (OC43 and HKU-1) or Alpha (NL63 and 229E) HCoV. Results were read as for ELISpot and presented as expansion compared to the DMSO controls.

### Cytokine measurement

Supernatants from donors with a detectable response in overnight ELISpot cultures were assessed using a LEGENDplex Th-profile 12-plex kit (Biolegend) following manufacturer’s instructions. Data was analysed using LEGENDplex v8.0 Software (Biolegend).

### Intracellular Cytokine Staining

Cryopreserved PBMC were thawed and rested overnight. Cells were then stimulated with a combined spike S1 and S2 peptide pool, at a final concentration of 1μg/ml per peptide, or DMSO (Mock). After 1hr eBioscience Protein transport inhibitor cocktail (Thermofisher Scientific) was added and cells incubated for a further 5 hrs. eBioscience Cell stimulation cocktail (Thermofisher Scientific) was used as a positive control. Following stimulation, cells were washed (PBS+0.1%BSA) and surface stained at 4°C for 30min. Cells were then washed and fixed in 2% paraformaldehyde. Following washing, brilliant staining buffer was added (BD Bioscience) and a final concentration of 0.4% Saponin added. Cells were stained intracellularly at room temperature for 30min. Cells were then washed and run on a BD Symphony A3 flow cytometer (BD Biosciences). Antibody details are provided in Extended Method Table 1.

### Data visualisation and statistics

Statistical tests, including normality tests, were performed as indicated using GraphPad Prism v9 software. Only results found to be significant (p<0.05) are displayed.

## Results

### Children develop robust and broad antibody responses after SARS-CoV-2 infection

Blood samples were obtained from 91 children and 154 adults, including 35 children and 81 adults known to be seropositive in previous rounds of testing. All infections were asymptomatic or mild and no staff or students in the cohort required medical care or hospitalisation. The median age of the children was 7 years (range, 3-11) whilst that of adults was 41 years (range, 20-71). SARS-CoV-2 antibody profile was assessed using the MSD V-plex serology platform to determine serological responses against spike, Receptor Binding Domain (RBD), N-terminal domain (NTD) and Nucleocapsid (N). In total, 47% of children and 59% of adults were found to be seropositive (Extended Data Table 1). To ensure the sensitivity of our assays we obtained convalescent plasma samples from 35 SARS-CoV-2 PCR-confirmed children. 34 of these were seropositive in the assay whilst one donor mounted no detectable antibody response to any antigen tested. Pre-pandemic plasma samples from 9 children and 50 adults all gave negative results and demonstrate specificity of the assay (Extended Data Figure 1).

Seropositive children and adults demonstrated broadly similar antibody responses against viral proteins. Geometric mean antibody titres against all four regions were, however, higher in children, most notably against the NTD and RBD domains, which showed 2.3-fold and 1.7-fold increases respectively, although these did not reach statistical significance. (Figure 1A). In contrast to previous reports ^3^ we also observed antibody responses against nucleoprotein, with a 1.3-fold increased antibody titre compared to adults (Figure 1 A&B).

**Figure 1.**
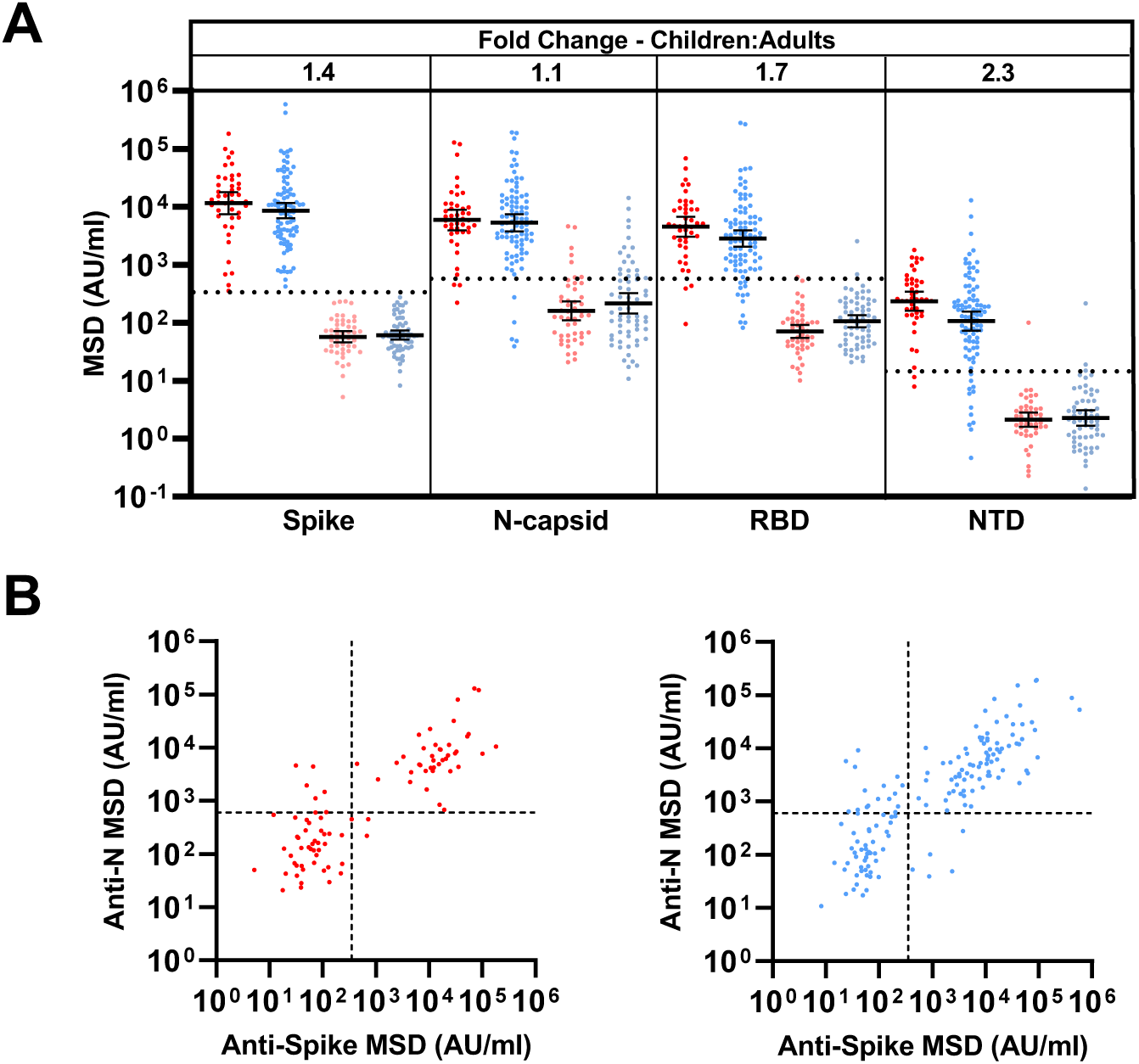
Children and adults develop coordinated antibody responses to SARS-CoV-2. A) SARS-CoV-2 antibody levels determined by MSD assay in children (n=91) and adults (n=154). Serostatus was determined based upon spike serology and used to divide the cohorts into seropositive (red/blue) and seronegative (light red/light blue). Seropositive/negative children n=43/48; adults n=91/63 respectively. Dotted lines represent cut off values for serostatus. Fold-change indicates the difference between the geometric mean titres in seropositive children and adults. Bars indicate geometric mean, with 95% CI. **B)** The level of spike-specific and nucleocapsid-specific antibody response was correlated within individual donors and reveals a coordinated response to both proteins.

### SARS-CoV-2 infection boosts antibodies responses against HCoV in children

Pre-existing immune responses against seasonal coronaviruses might act to modulate clinical outcome following primary SARS-CoV-2 infection, and cross-reactive neutralising antibodies have been reported in SARS-CoV-2-seronegative children^15^. Consequently, we compared antibody levels against the four HCoV in SARS-CoV-2 in seronegative and seropositive children and adults.

A 1.2 to 1.4-fold increase in the titre against HCoV was evident in SARS-CoV-2 seropositive adults compared to the seronegative group. In contrast, antibody levels against all four viruses were boosted markedly in SARS-CoV-2 seropositive children, with 2.3, 1.9, 1.5, and 2.1-fold higher antibody levels compared to the seronegative group. These were significant for OC43 and HKU-1 (p=0.0071 and p=0.0024 Brown-Forsythe and Welch ANOVA, with Dunnett’s T3 multiple comparison test) (Figure 2A). Notably, the level of HCoV-specific antibodies in seropositive children was comparable to adults, whereas seronegative children possess lower responses than adults (Extended Data Table 1).

**Figure 2.**
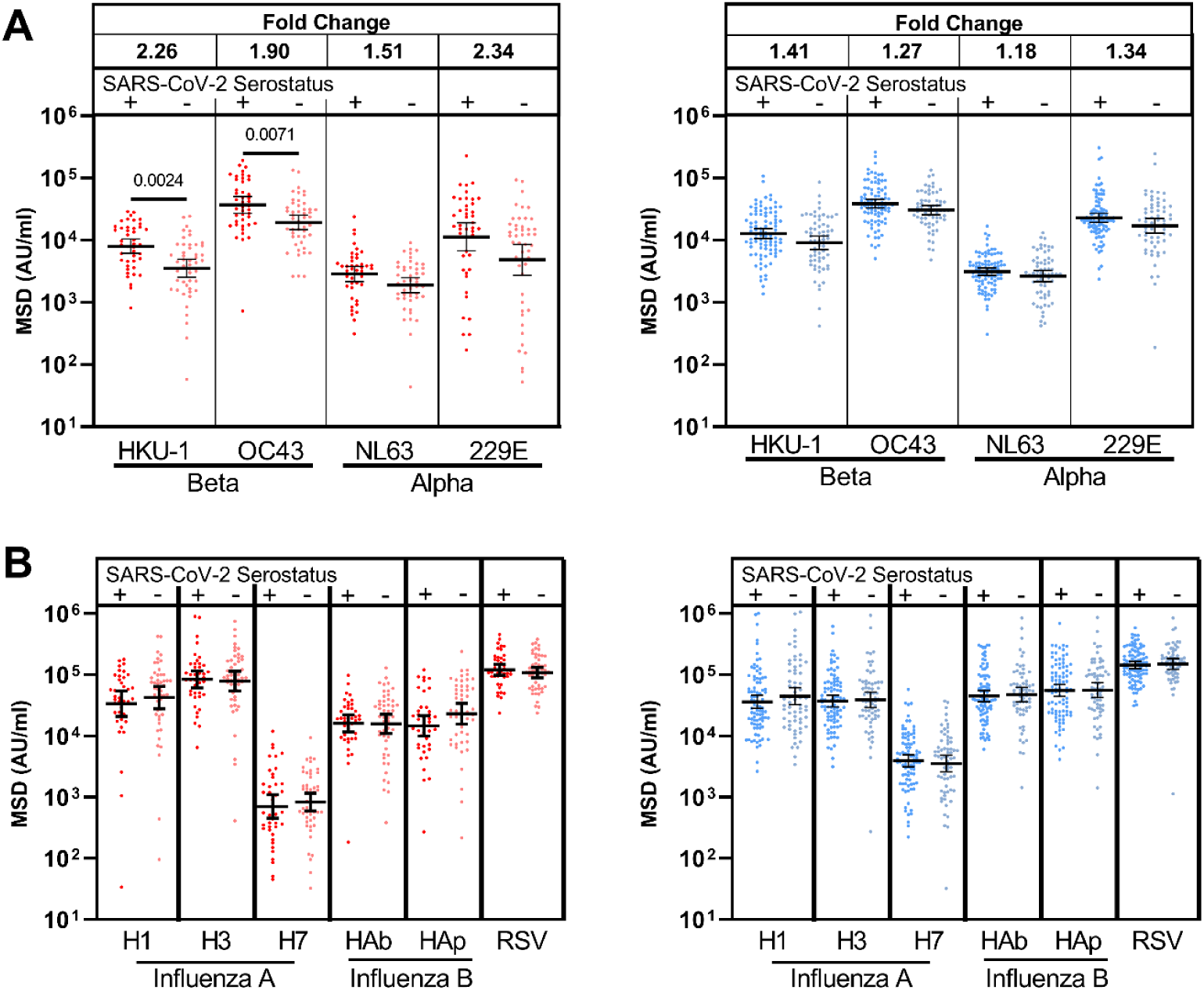
Antibody responses against the seasonal HCoV Beta-coronaviruses are back-boosted in children following SARS-CoV-2 infection. (A) Antibody titres to the seasonal HCoV coronaviruses and (B) other respiratory viruses in children (red) and adults (blue) based on SARS-CoV-2 serostatus (dark=seropositive, light=seronegative). Seropositive/negative children n=43/48; adults n=91/63 respectively. Fold change indicates the difference between the geometric mean titres in seropositive children and adults. Bars indicate geometric mean with 95% CI. Only significant differences are shown. Brown-Forsythe and Welch ANOVA, with Dunnett’s T3 multiple comparison test.

In order to assess if this effect was specific to HCoV or represented a more general effect of SARS-CoV-2 infection on antibody responses against heterologous infection, we also examined antibody titres against influenza subtypes and respiratory syncytial virus (RSV) in relation to SARS-CoV-2 serostatus. No significant change in antibody titre against these viruses was seen in either children or adults (Figure 2B). These data show that SARS-CoV-2 infection in children specifically boosts humoral responses against HCoV.

### SARS-CoV-2-specific antibodies in children can cross-react with the S2 domain of Beta-coronaviruses

Given the increase in HCoV-specific antibody titres following SARS-CoV-2 infection in children we next assessed to what extent this was cross-reactive against SARS-CoV-2 or could represent a HCoV-specific response. As such, recombinant S1 or S2 domain protein from SARS-CoV-2 was used to pre-absorb plasma samples prior to assessment of antibody levels to both SARS-CoV-2 and the four HCoV subtypes.

As expected, pre-absorption with both the S1 and S2 domains significantly reduced antibody titres against total spike (p<0.0001 and p=0.0024, respectively, Friedman test with Dunn’s multiple comparisons test). The S1 domain but not the S2 domain, absorbed RBD and NTD-specific antibodies against SARS-CoV-2 whilst no influence was observed in relation to nucleocapsid-specific binding for either domain (Figure 3). Of note, the S1 domain did not significantly reduce antibody binding to any of the 4 HCoV subtypes indicating little evidence for cross-reactive antibodies against this domain. The S2 domain, however, selectively reduced antibody binding to the two HCoV Beta-coronaviruses, OC43 and HKU-1 (p <0.0001 and p=0.0014, respectively by repeated measure one-way ANOVA with Holm-Sidak’s multiple comparison test). No such effect was observed in relation to binding to the Alpha-coronaviruses NL63 and 229e (Figure 3).

**Figure 3.**
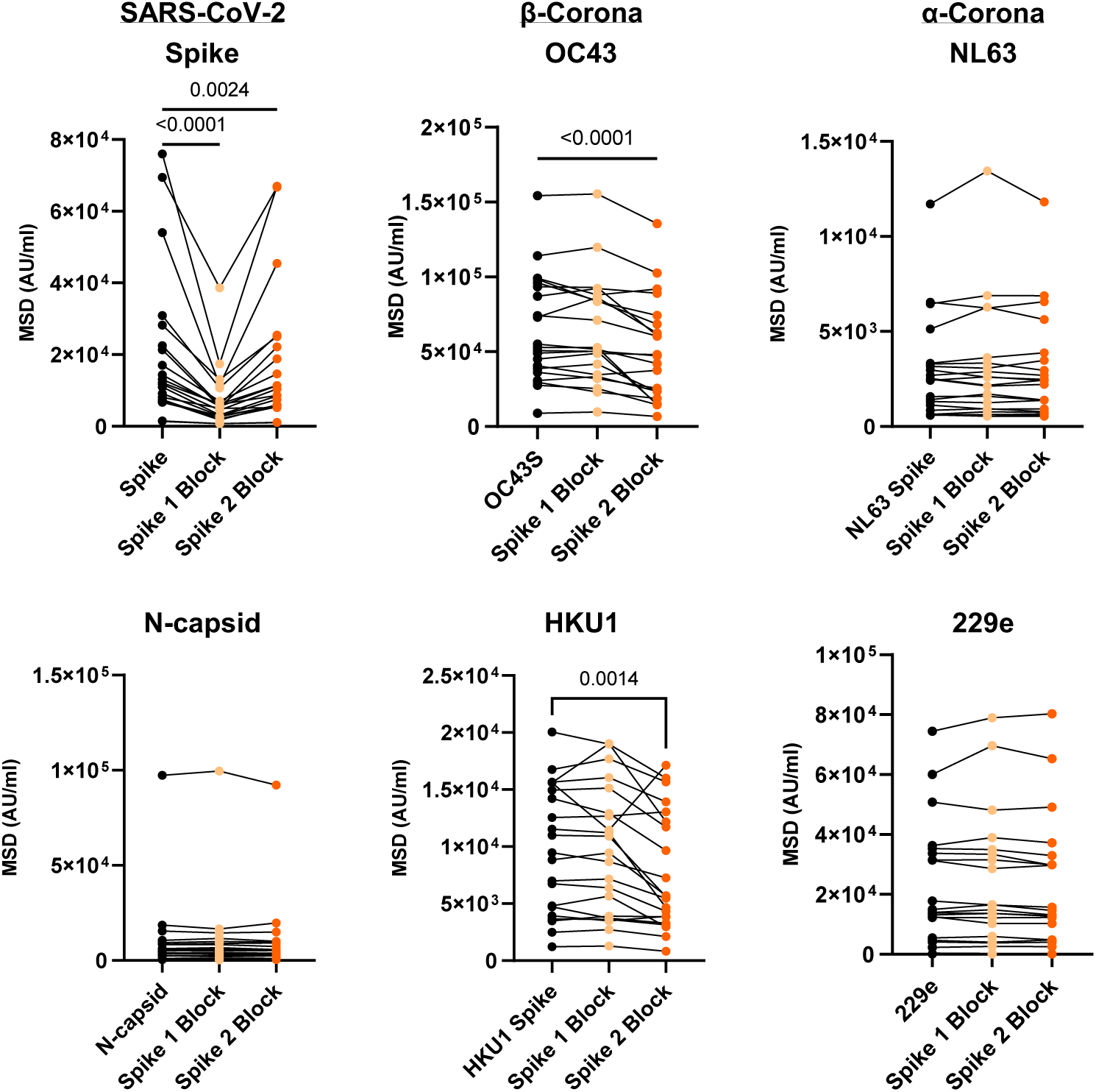
Cross reactive antibodies against the spike S2 domain contribute to increased HCoV-specific antibody responses in SARS-CoV-2 seropositive children. (A) Plasma from SARS-CoV-2 seropositive children (n=21) was assessed for binding to the spike protein of the four HCoV or the spike or nucleocapsid regions of SARS-CoV-2. Plasma was either applied neat (control) or following pre-absorption with either recombinant spike S1 domain (spike 1 Block) or spike S2 domain (spike 2 Block). S1 pre-absorption markedly reduced binding to SARS-CoV-2 spike whilst S2 pre-absorption reduced binding to OC43 and HKU-1. Repeated measure one-way ANOVA with Holm-Sidak’s multiple comparison test, or Friedman test with Dunn’s multiple comparisons test as appropriate

These data show that the S1 region is the immunodominant target of antibody responses in children. However, antibodies that are cross-reactive against Beta-coronavirus are largely specific for the S2-domain and contribute to the higher SARS-CoV-2-specific titre in children.

### Children develop robust cellular immune responses against spike protein after SARS-CoV-2 infection

We next assessed the magnitude and profile of the cellular immune response against SARS-CoV-2 in children and adults. ELISpot analysis against overlapping peptide pools from spike and a combination of nucleocapsid and membrane and envelope (N/M) was performed on samples from 57 children and 93 adults, including 37 and 64 respectively who were seropositive.

As expected, ELISpot responses were common in SARS-CoV-2 seropositive donors with 89% (33/37) of seropositive children and 80% (51/64) of seropositive adults showing a positive ELISpot response to spike and/or the N/M pool.

The magnitude of the cellular response against spike was 2.1-fold higher in children, with median values of 533 spots/million compared to 195 in adults (p=0.0003, Brown-Forsythe and Welch Anova, with Dunnett’s T 3 multiple comparisons test) (Figure 4A). Cellular responses against the N/M pool were relatively lower in children compared to spike such that the S:N/M ratio was markedly elevated in children at 4.7 compared to 1.8 in adults (p=0.0007, Brown-Forsythe and Welch Anova, with Dunnett’s T 3 multiple comparisons test) (Figure 4B). 86% of children showed a positive response to spike whilst only 43% responded to the N/M pool. Within adults these values were 70% and 63% respectively (Figure 4C).

**Figure 4.**
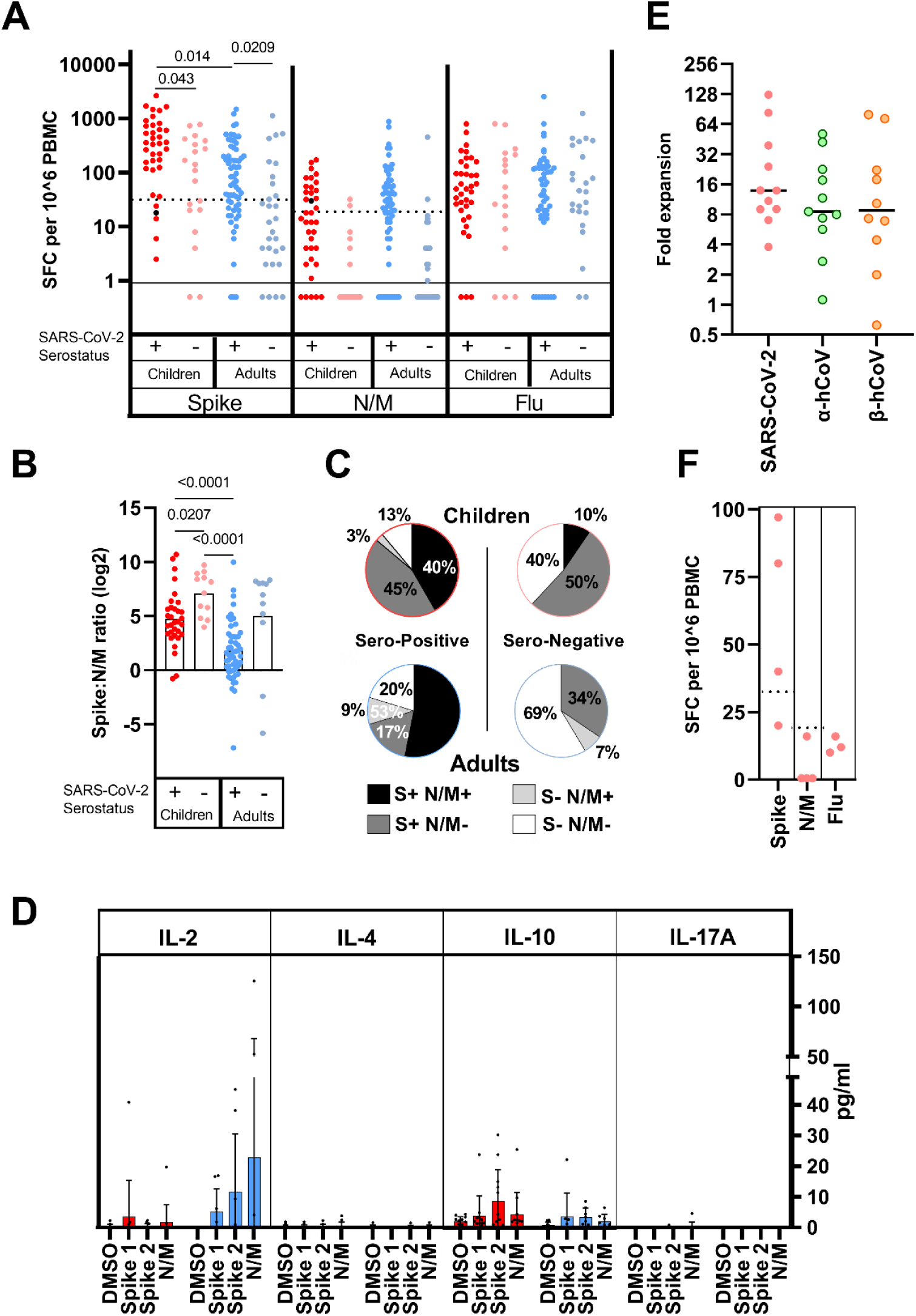
Children develop strong spike-specific T cell responses after SARS-CoV-2 infection and cross-reactive responses are also found in many seronegative donors. A) SARS-CoV-2-specific T cell responses in children (n=57; red) and adults (n=83; blue). SARS-CoV-2 serostatus was 37/20 seropositive or negative in children and 64/29 seropositive or negative in adults respectively. Assay used IFN-γ ELISpot using pepmixes containing overlapping peptides to spike, Nucleoprotein and viral Membrane (N/M), or Influenza, and is shown in relation to serostatus. B) The magnitude of spike-specific cellular response was compared to that against N/M and displayed as a ratio in seropositive and seronegative adults and children, as indicated. Bars indicate mean, Brown-Forsythe and Welch Anova, with Dunnett’s T 3 multiple comparisons test. C) The proportions of subjects within each cohort which demonstrate a cellular response to S or N/M peptides from SARS-CoV-2. D) Cytokine concentration within supernatants from ELISpot cultures (n= 12 children (red); n=8 adults (blue)). Bars indicate mean ±SD. E) HCoV-specific cellular responses show equivalent expansion following stimulation of PBMC from SARS-CoV-2 seronegative children with SARS-CoV-2 S2 domain pepmix (n=11). Cultures were stimulated for 9 days and then assessed by IFN-γ ELISpot to pepmix of S2 domain from SARS-CoV-2 or the alpha (OC43 and HKU-1) or beta (NL63 and 229E) HCoV. Expansion is shown relative to unstimulated control cultures. Lines indicate median. F) SARS-CoV-2-specific T cell response in PBMC samples taken from children prior to the CoVID-19 pandemic (n=4).

Supernatant from ELISpot was analysed using a multi-analyte bead assay to compare the profile of cytokine production by SARS-CoV-2-specific T cells from children and adults. Samples from adult donors showed a marked IL-2 response with lower levels of IL-10 production. In contrast, IL-2 levels in samples from children were very low (Figure 4D) indicating differential functional response compared with adults. Indeed, analysis of three children by flow cytometry indicated that CD8+ IL-2-TNF+IFN-γ+ T cells constituted the bulk of the spike-specific T cell response in children (Extended Data Figure 2).

Notably, cellular responses were also observed in 60% (12/20) of seronegative children, all of whom were seronegative by three different serology platforms. Cellular responses of variable but lower magnitude were also present in 34% (10/29) of seronegative adults (Figure 4 A). Two children and one adult classed as seronegative had anti-nucleocapsid antibodies (Extended Data Table 2.) but this was considered insufficient to provide definitive serostatus. These cellular responses in seronegative donors were markedly spike-specific, with elevated S:N/M ratios of 7.1 and 5 in children and adults respectively (Figure 4B), indicating pre-existing cross-reactive immunity. Indeed seronegative children (7/12) and seronegative adults (6/10) with a positive ELISpot demonstrated high antibody levels to one or more HCoV (Extended Data Table 2), potentially indicating recent HCoV infection.

To examine the presence of cross-reactive T cells in seronegative children we hypothesised that expansion of T cells in response to SARS-CoV-2 would be associated with an associated increase in HCoV-reactive responses. As such we first stimulated cells from SARS-CoV-2 seronegative donors with SARS-CoV-2 peptides and then assessed the response to peptides from SARS-CoV-2 and also peptide pools from the Alpha and Beta HCoV. Markedly increased cellular responses to both Alpha and Beta HCoV were seen after culture (8.6-fold and 8.9-fold respectively) (Figure 4E), indicating SARS-CoV-2-driven expansion of broadly cross-reactive T cells. Finally, to definitively assess the presence of cross-reactive T cells in children we obtained pre-pandemic PBMC from children and observed that 50% of these had notable responses to spike by ELISpot but lacked cellular responses against N/M peptides. Matched plasma samples, available for three donors, showed that antibody responses against HKU-1 and 229E were 2 and 5-fold greater in a donor with high cellular responses compared to two children that lacked cellular responses.

Overall these data show that cross-reactive coronavirus-specific T cell responses are present within a high proportion of children.

### Children maintain immune responses against SARS-CoV-2 for at least 12 months

We were next interested to assess the longevity of immune responses within a subgroup of 35 children and 81 adults who had seroconverted at least 6 months prior to analysis. All children retained humoral immunity whilst 7% (6/81) of previously seropositive adults failed to show significant antibody responses. Children also maintained higher antibody titres against spike and RBD which were 1.8-fold higher than adults (Figure 5A&B).

**Figure 5.**
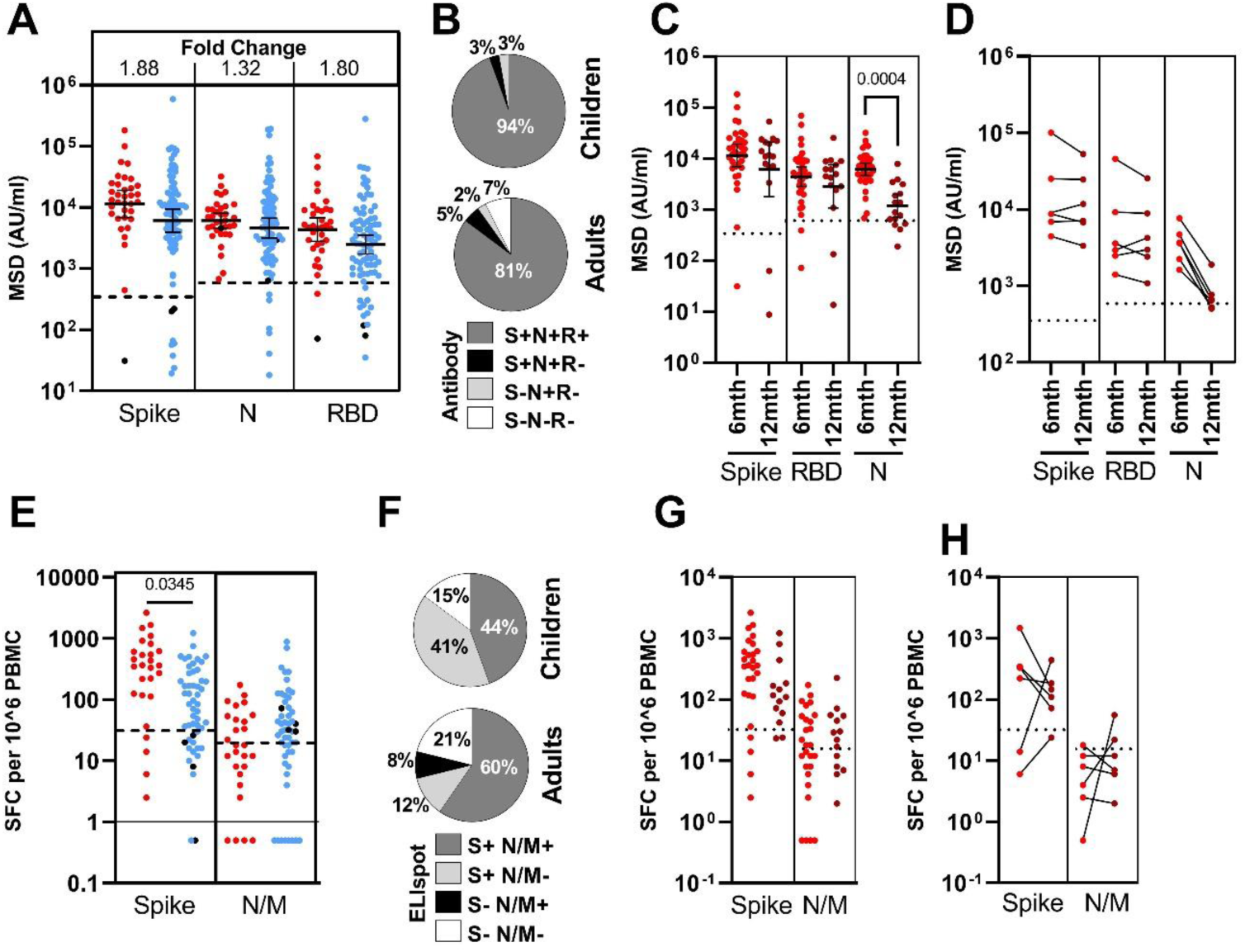
Higher spike-specific immune responses are maintained in children for at least six months after infection. A) Antibody responses in children (n=35; red) and adults (n=81; blue) who were seropositive at first testing and therefore at least 6-months post-primary infection. Bars indicate geometric mean titre (GMT) ± 95%CI. Dotted lines indicate seropositive cut-offs. Fold change indicates increment in children’s samples GMT compared to adults. Black dots indicate individuals who were seronegative for spike but retained a nucleocapsid-specific antibody response. B) Proportion of individuals retaining antibody responses to spike, Nucleocapsid or RBD at ≥6 months. C) Antibody binding to spike, RBD or N in children ≥6 months (n=35, red) or ≥12 months (n=16, dark red) after infection. Bars indicate GMT ±95%CI. Dotted lines indicate seropositive cut-offs. Kruskal-Wallis with Dunn’s multiple comparison test. D) Paired antibody levels for children (n=6) at ≥6 and ≥12 months post-primary infection. Dotted lines indicate positive cut-offs. E) Spike-specific IFN-γ ELISPOT in children (n=27; red) and adults (n=52; blue). Black dots indicate individuals who lacked a spike-specific response but retained a nucleocapsid-specific response. Brown-Forsythe and Welch Anova with Dunnett’s T 3 multiple comparisons test. F) The proportion of each cohort scored as responding to SARS-CoV-2 by ELISPOT. G) Spike-specific IFN-γ ELISPOT in children ≥6 months (n=27, red) or ≥12 months (n=14, dark red) after infection. Dotted lines indicate seropositive cut-offs. H) Paired ELISPOT results for children at ≥6 and ≥12 months post-primary infection (n=6). Dotted lines indicate positive cut-offs.

Samples were also obtained from 16 children who had seroconverted at least 12 months prior to analysis. Antibody levels to spike and RBD were retained at a similar, although slightly reduced, level to those seen at 6 months whilst nucleocapsid-specific antibody levels were significantly reduced (p=0.0004) (Figure 5C). 2 of these 16 children (12.5%) had spike-specific antibody levels below threshold whilst 4 children (25%) showed similar loss of nucleocapsid-specific antibodies, one of whom also lost spike-specific response. Six of these donors had been analysed previously at 6 months and matched individual comparisons revealed stable spike-specific antibody levels between 6 and 12 months post infection whereas nucleocapsid-specific antibodies showed significant waning, with 2 donors dropping below cut off (Figure 5D).

Cellular immune responses were detectable in 84% of children and 79% of adults at least 6 months after infection. The magnitude of the spike-specific response remained higher in children than in adults (p=0.032) whereas responses to the N/M pool were seen in only 31% of children compared to 68% of adults (Figure 5D&E). 15 children who had seroconverted over 12 months previously were also assessed by ELISpot. In comparison to cohort analysis at 6 months, T cell responses to spike were retained but somewhat reduced, whilst nucleocapsid-specific responses, although of lower magnitude, were retained at a similar level (Figure 5G). Matched samples at 6 and 12 months were available for 5 of these children and were stable. These data show that children broadly retain both antibody responses and cellular responses for extended periods following primary infection (Figure 5H).

### Antibodies from children show enhanced binding to spike protein from viral variants of concern but equivalent levels of viral neutralisation

SARS-CoV-2 variants of concern (VOC) may be able to partially escape immunity generated by prior infection or vaccination ^22, 23^. Given the development and maintenance of cross-reactive high-level antibody responses in children, we were interested to assess their relative recognition of spike protein from VOC. Plasma samples from 19 children and 18 adults at >6 months after primary infection were tested for binding to spike and RBD from alpha (B.1.1.7), beta (B.1.351) and gamma (P.1) variants compared to the original Wuhan genotype, which was used in previous assays.

As described above, children, compared to adults, maintain higher levels of antibody binding to Wuhan spike (Figure 5A, Figure 6A) and this was also observed in binding to spike from the three VOC with 1.7, 1.8, and 2.1-fold higher geometric mean titres (GMT) against alpha, beta and gamma variants, respectively (Figure 6A). Children also demonstrated higher binding to the RBD region of the three VOC, compared to adults, with 2.1, 1.8 and 2.9-fold higher GMT, respectively (p=0.029 and p=0.0114 against beta and gamma, respectively; Kruskal-Wallis test with Dunn’s multiple comparison test). Applying the threshold determined in earlier assays, 16/19 (84%) children retained seropositive status to both beta and gamma compared to only 5/18 (28%) and 8/18 (44%) of adults, respectively (Figure 6B). Similar ratios of relative binding or inhibition of spike-ACE2 engagement with spike from Wuhan or VOC were seen in children and adults (Extended Data Figure 3 A&B, Extended Data Table 3) indicating that enhanced antibody binding to VOC is a function of overall quantitively superior antibody responses in children.

**Figure 6.**
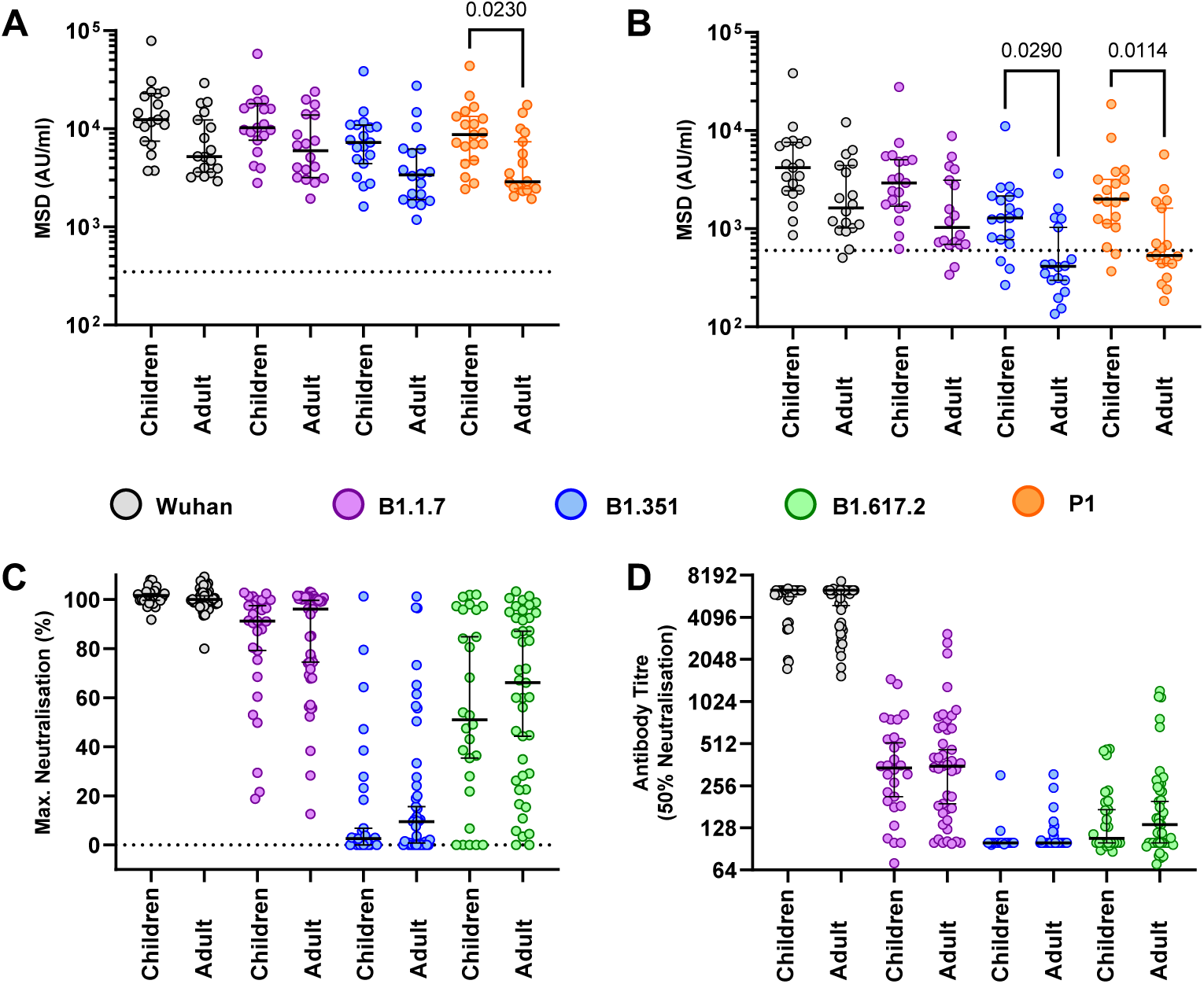
Plasma from children at 6+ months after infection shows superior antibody binding to spike and RBD from SARS-CoV-2 viral variants but comparable neutralisation of live virus. (A) Antibody binding to spike and (B) RBD protein from SARS-CoV-2 variants using plasma from children (n=19) or 18 adults (n=18). Bars indicate Geometric Mean±95%CI. Kruskal-Wallis test with Dunn’s multiple comparison test. (C) Live virus neutralisation assays on SARS-CoV-2 variants displayed as Maximal neutralisation of infection and (D) titre at 50% neutralisation using plasma from children (n=28) or adults (n=43).

**Figure 7.**
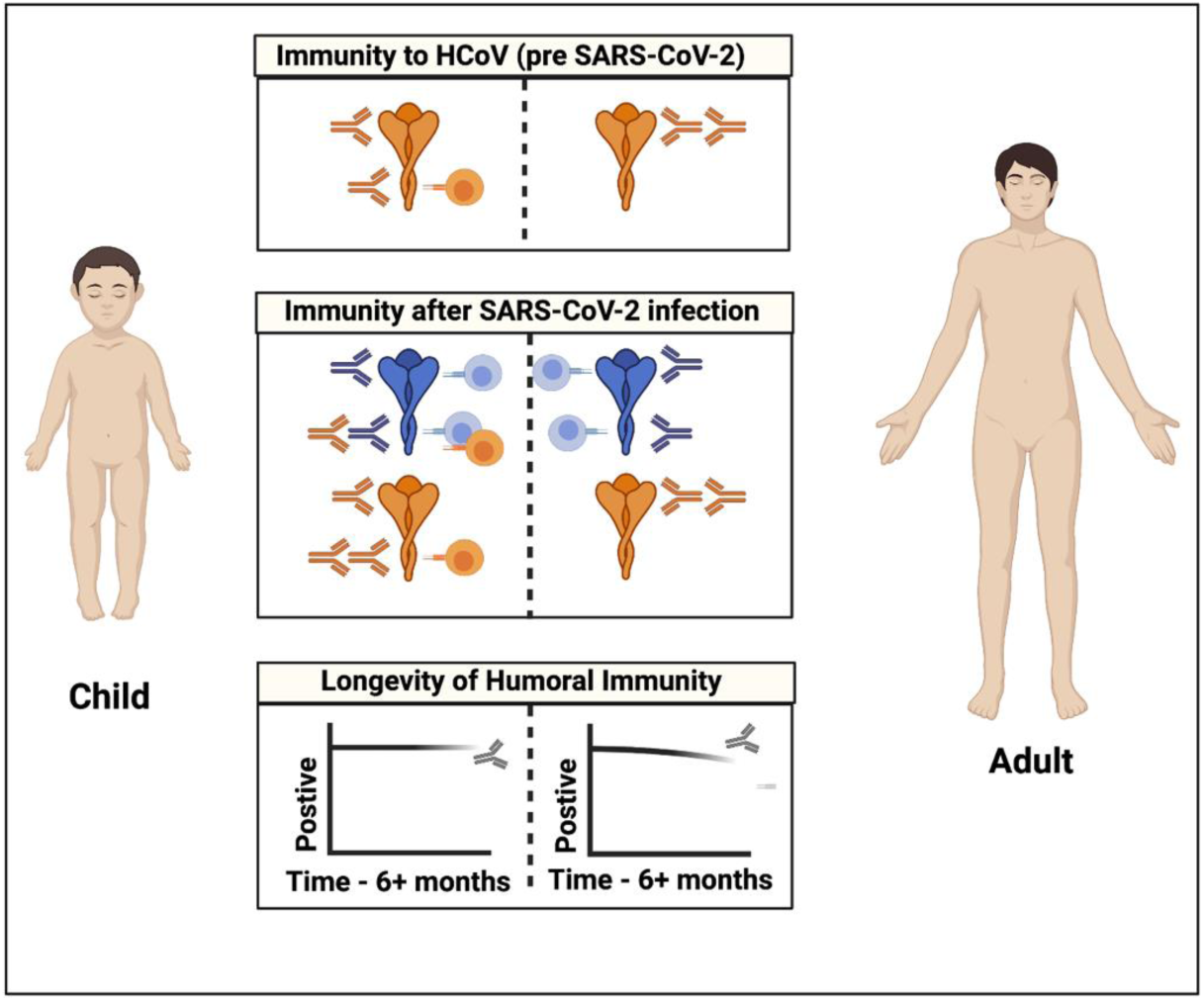
Model of antibody and cellular immunity to seasonal human coronaviruses (HCoV) and SARS-CoV-2 in children and adults. Children develop HCoV spike S2-specific antibody and cellular responses that can cross react with SARS-CoV-2. Robust S1-specific adaptive responses develop following SARS-CoV-2 infection. Cross reactive S2-specific responses likely contribute to immune control in children. Adapted from “Coronavirus Replication Cycle”, by BioRender.com (2020). Retrieved from https://app.biorender.com/biorender-templates

We next assessed the ability of sera from children and adults to neutralise infection by live virus. Serum from adults showed markedly reduced capacity to neutralise the VOC and this was particularly noteworthy for B.1.351 (Figure 6C&D). This pattern was also seen in children and no difference was seen in either maximal or 50% neutralisation titre between adults and children. A similar profile was seen with a pseudotype-based neutralisation assay (Extended Data Figure 3D).

These data show that children develop higher antibody binding to SARS-CoV-2 VOC after natural infection compared to adults but display similar neutralising ability.

## Discussion

Age is the primary determinant of the clinical severity of SARS-CoV-2 infection and a life course assessment of virus-specific immunity is essential to understand disease pathogenesis and design vaccine strategies in children. Our detailed analysis of adaptive immune memory identifies a number of novel features in young children. A key finding was that the magnitude of the adaptive immune response to SARS-CoV-2 is higher in children compared to adults. This is somewhat different to previous reports which have reported lower T cell responses in children^6^. This may reflect differences in assay systems as here we used separate spike and N/M peptide pools to demonstrate the heightened spike-specific response. It has also been reported that children do not mount effective antibody responses against nucleocapsid in the early post-infection period ^3, 5^. Using the well validated Meso Scale Diagnostics system we did observe nucleocapsid-specific antibody responses in children but it was noteworthy that immune responses were much more focussed against spike ^24^. Nucleocapsid is an abundant protein within the SARS-CoV-2 virion and it is possible that the magnitude of the N-specific response is a reflection of peak or aggregate viral load. Levels of virus within the upper airways at the time of primary infection are equivalent in children and adults ^25^ but relative changes over the course of infection are not known. Enhanced innate immune responses in children may also play an important role in limiting systemic replication and may explain the higher rates of asymptomatic and mild illness in children compared to adults ^26^. Antibody levels generally correlate with disease severity but none of the children or adults in this study had suffered from severe disease or needed hospital admission

One striking feature was that SARS-CoV-2 infection in children doubled antibody titres against all four Alpha and Beta HCoV subtypes. This pattern was not seen in adults where increased humoral responses were modest ^27, 28^. Of note, increased antibody titres against HCoV have also been observed after SARS-CoV-1 infection ^29^. Using protein domain pre-absorption we find that most of this response results from SARS-CoV-2-specific humoral responses that cross-react with the S2 domain of the two more closely related Beta-coronaviruses. The S2 domain is more highly conserved between HCoV than S1 and this pattern is compatible with preferential targeting of structurally-conserved epitopes by HCoV-specific antibodies in children ^30^ with the potential for neutralising activity against SARS-CoV-2 ^15, 31^. However, SARS-CoV-2 infection in children also boosts HCoV-specific antibody responses that are not directly cross-reactive, as demonstrated by increased titres against Alpha-coronaviruses that could not be pre-absorbed. This may potentially reflect weakly cross-reactive B cell clones, potentially activated through T cell cross-recognition, and is reminiscent of antibody boosting against H3 haemagglutinin after H1N1 infection in children who have had previous H3N2 infection ^32^. We found that titres of HCoV-specific antibodies are lower in seronegative children in comparison to adults but these are likely to increase following primary infection during late teenage years ^13, 30^ and it appears that repeated infections may hone S1 domain-specific responses and lead to loss of cross-reactive S2-specific responses in adulthood ^15^. Our data show that SARS-CoV-2 infection acts to fill the CoV-specific antibody space, producing antibodies against all coronaviruses and potentially supporting immunity in later life.

The development of antibodies against Alpha-coronaviruses that are not absorbed by the S2 domain of SARS-CoV-2 suggests that there may be a lower affinity threshold for boosting of related but heterologous immune responses in children which might potentially represent an evolutionary adaptation to expand the memory pool early in life. It is interesting to speculate whether this may provide insight into the pathophysiology of PIMS-TS/ MIS-C, where B cell activation drives a hyper-inflammatory syndrome. Lack of pre-existing immunity to HCoV is a risk factor for PIMS-TS/MIS-C and may indicate a protective influence from a prior cross-reactive memory B cell pool ^8^. Furthermore, the pathogenic antibodies associated with this condition are enriched for antibodies against heterologous pathogens and inflammation may result from FcR-mediated monocyte activation ^33^.

This profile of an enhanced and cross-reactive humoral immune response in children was also apparent in analysis of the cellular response to SARS-CoV-2. We find that the virus-specific T cell response is higher in children compared to adults and this mirrored the humoral response in that responses against the spike protein were markedly increased compared to nucleocapsid and envelope proteins. Virus-specific T cell responses in children also showed a differential cytokine response with markedly reduced production of IL-2. This may suggest a more highly differentiated profile in children compared to adults, in line with the enhanced magnitude of the response. Indeed, the spike-specific CD8+ T cell pool in children at 6 months after primary infection was dominated by an IL-2^-^IFN-γ^+^TNF^+^ phenotype. As such further long-term characterisation of the T cell response in children, and the potential mechanisms that may act to drive T cell activation, are required.

We also observed that cross-reactive CoV-specific T cells are present in many children, both before and after SARS-CoV-2 infection. SARS-CoV-2-specific T cell responses were detectable in more than half the seronegative children, including samples taken pre-pandemic, and are likely to represent HCoV-specific T cell responses that cross react against SARS-CoV-2 peptides ^34, 35^. Of note, young adults have been recently reported to have higher T cell responses to HCoV than older people and these can cross-react with SARS-CoV-2 ^36^. SARS-CoV-2 -reactive T cells were also seen in some seronegative adults and it is also possible that some of these responses represent genuine SARS-CoV-2-specific T cells that have been generated following virus exposure in the absence of antibody sero-conversion. This pattern has been reported in health care workers with high levels of viral exposure ^37^ and it is possible that such conditions are also seen in primary schools where enforcement of social distancing is challenging.

It is tempting to speculate that this profile of cross-reactive antibody and cellular responses in children contributes to the excellent clinical outcomes in this group. In this regard, it would be interesting to assess matched paediatric samples from before and after SARS-CoV-2 infection in order to determine the relative contribution of cross-reactive HCoV-specific clones to the protective adaptive immune response.

Many participants were seropositive 6 months prior to analysis and our data extend previous findings in adults ^21, 38^ to show sustained immunity over this time period. Moreover, we also found that children maintained higher antibody and cellular immune responses at this timepoint compared to adults with no loss of humoral response compared to 7% in adults. There is increasing confidence in the relative stability of SARS-CoV-2-specific memory B cell and antibody responses but studies of antibody waning after natural infection are now difficult to perform in adults due to the widespread adoption of Covid-19 vaccines. Spike-specific antibody responses were also largely maintained in children at 12 months, whereas responses against nucleocapsid showed significant waning. T cell responses were broadly stable and it is important to consider that ELISpot analysis will only detect effector populations and a long-term memory subset is likely to have been established by this point. We were not able to compare 12-month values to the adult population as this latter group had undergone COVID-19 vaccination prior to this timepoint.

We also found that children possess enhanced binding of antibodies to spike and RBD from viral VOC ^23^. However, levels of neutralising antibodies in live and pseudo-virus neutralisation assays were comparable between adults and children. This suggests that children and adults develop comparable neutralising antibody responses against the spike S1 domain but the increased antibody response in children results from antibodies targeting non-neutralising epitopes within S1 and S2. These antibodies could still have important effector potential through mechanisms such as antibody-directed cell cytotoxicity and further studies to examine the specificity and function of the SARS-CoV-2 B cell repertoire in children following natural infection or vaccination would be of value. In the light of the concern that SARS-CoV-2 will become an endemic infection ^39^, these findings augur well for immunity generated in childhood providing robust and sustained protection, including to emerging VOC.

In conclusion, we show that children display a characteristic robust and sustained adaptive immune response against SARS-CoV-2 with substantial cross-reactivity against other HCoV. This is likely to contribute to the relative clinical protection in this age group but these findings may also provide insight into the characteristic immuno-pathology that may develop. Furthermore, they will help to guide the introduction and interpretation of vaccine deployment in the paediatric population.

## Data Availability

Data is available following reasonable requests.

## Data Availability

The data that support the findings of this study are available from the corresponding author upon reasonable request.

## Acknowledgement

This work was partly funded by UKRI/NIHR through the UK Coronavirus Immunology Consortium (UK-CIC). We acknowledge the support of the G2P-UK National Virology Consortium (MR/W005611/1) funded by the UKRI. The study was also funded in part by the MRC (MC UU 1201412).

We would like to express our gratitude to the staff, parents and especially children for their participation in this study, without whom this work would not have been possible.

---

**Extended Data Figure 1.**
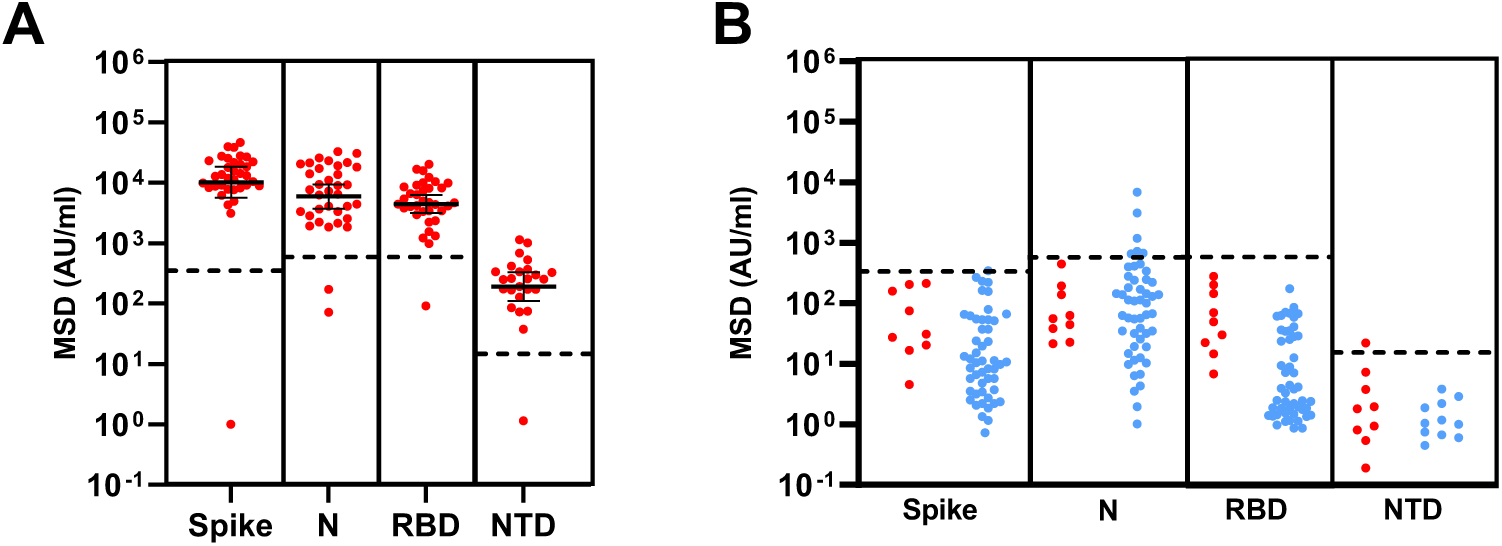
MSD assay specificity and sensitivity. A) MSD results using convalescent plasma samples from 35 children with PCR-confirmed SARS-CoV-2 infection. B) Assay cut-offs were tested using plasma samples from nine children (red) and 50 adults (blue) taken prior to COVID-19. Dotted lines indicate the cut-off used.

**Extended Data Figure 2.**
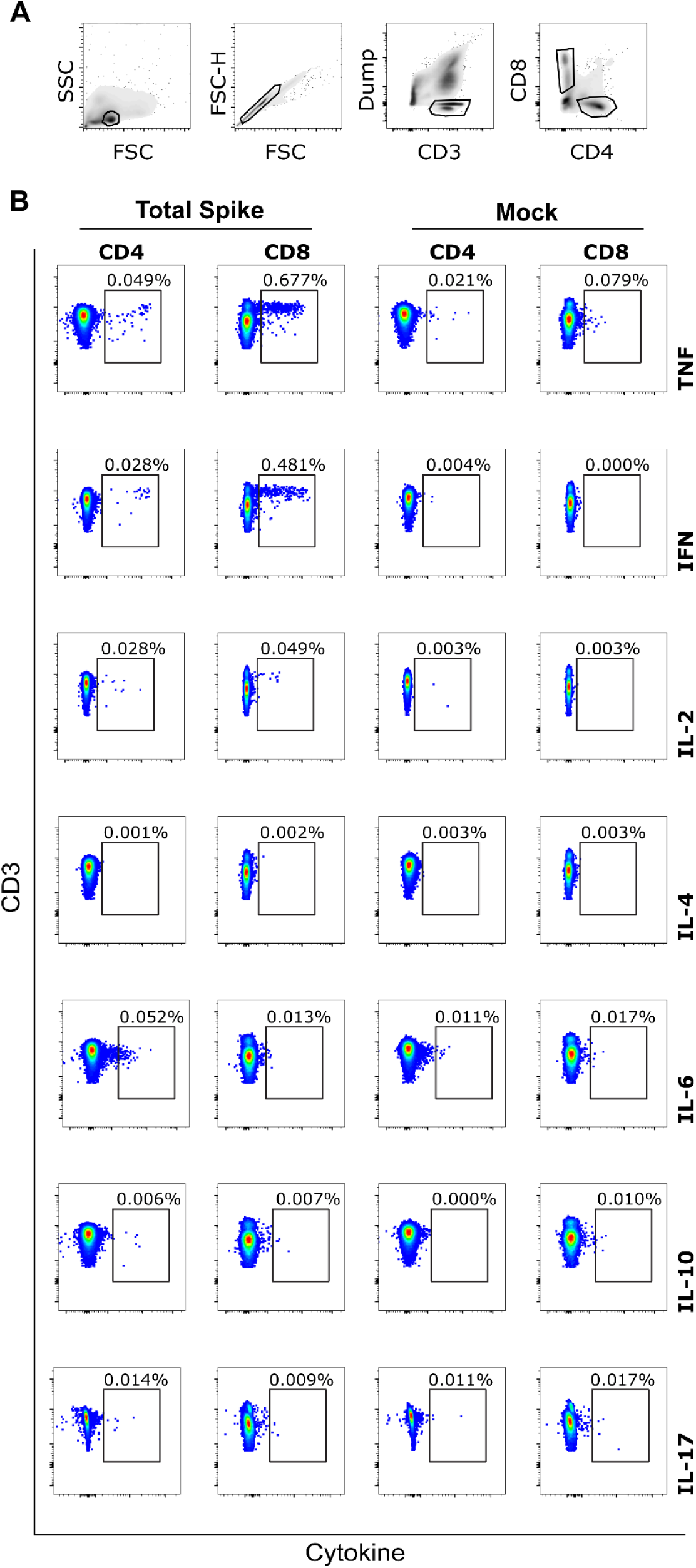
CD8+ T cells with a IL-2^-^TNF^+^IFN-γ^+^ phenotype dominate the spike-specific T cell response in children. PBMC from SARS-CoV-2 seropositive children were stimulated for 6hrs in the presence of spike peptide pool and then analysed by flow cytometry to assess intracellular cytokine stimulation (ICS). A) Gating strategy for analysis. B) Representative example of ICS staining from one child at six months post SARS-CoV-2 infection showing TNF+IFN-γ+ CD8+ T cell response representing 0.48% of the global CD8+ T cell repertoire.

**Extended Data Figure 3.**
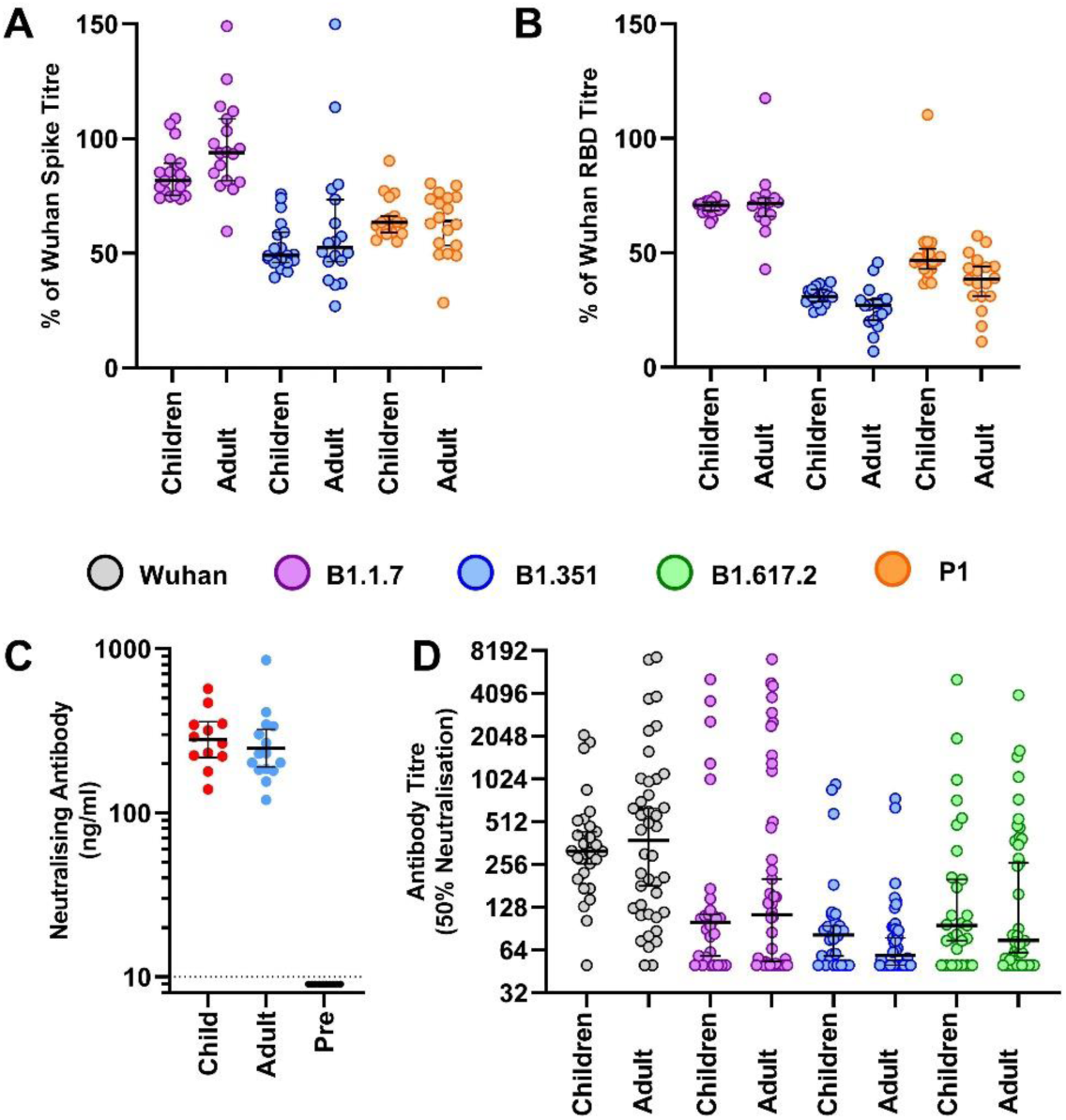
Antibody binding, RBD-ACE2 inhibition and pseudo-neutralisation of SARS-CoV-2 VOC in plasma from children or adults. (A) Antibody titres normalised to the Wuhan sequence as determined by MSD to total spike and (B) RBD from VOC as indicated in children (n=19) and adults (n=18) ≥6 months post primary infection. Bars indicate Median±95%CI. (C) Antibody inhibition in an RBD-ACE2 competitive binging assay using plasma ≥6 months post infection in children (n=12) and adults (n=15). Results from pre-pandemic adult samples are also shown (n=10). Bars indicate geometric mean ±95% CI. (D) Results from pseudo-virus neutralisation assays displayed as titre at 50% neutralisation from children (n=28) and adults (n=43) ≥6 months post primary infection.

**Extended Data Table 1.**
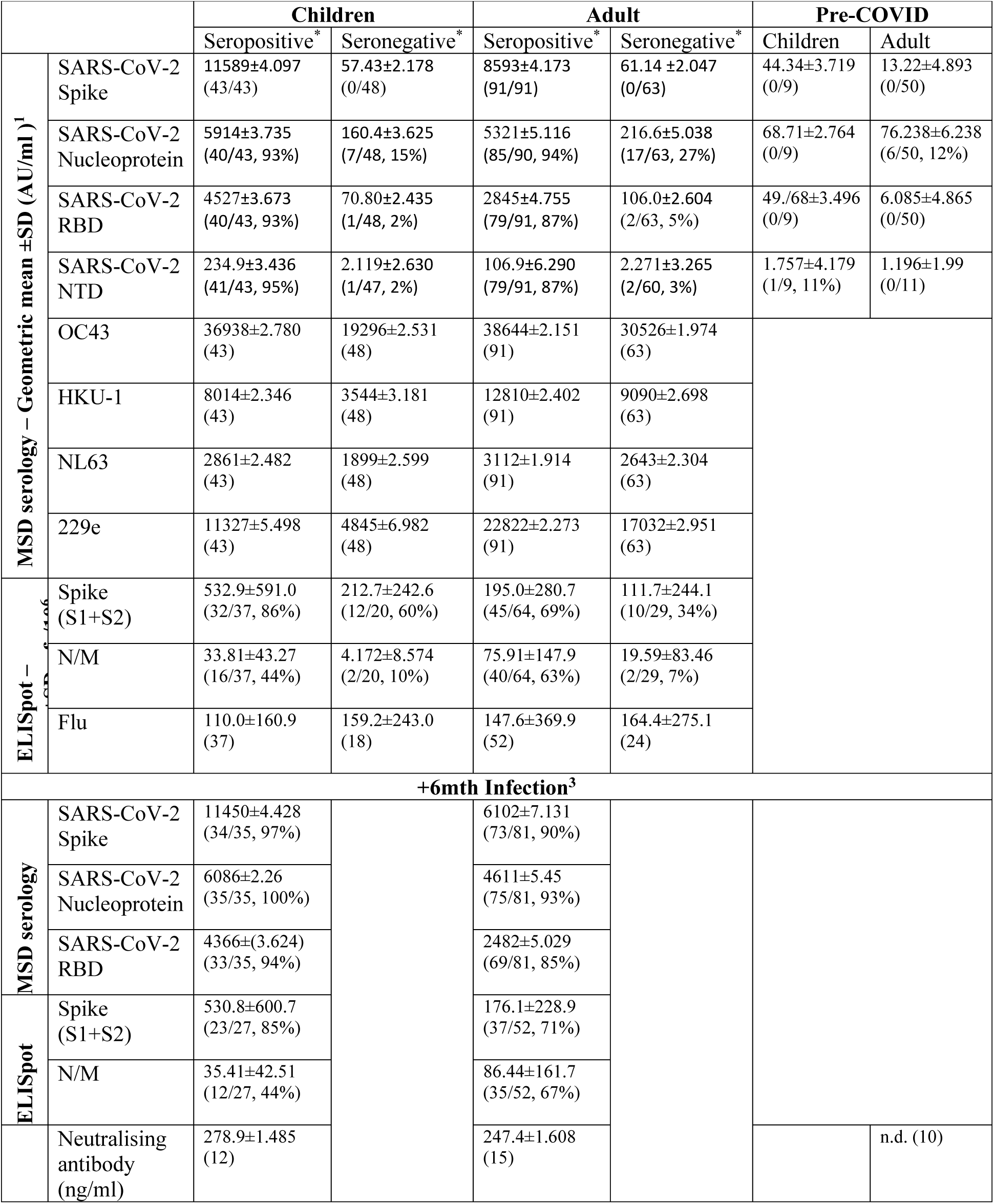
Immunological Characteristics of Seropositive and Seronegative Children and Adults. *Seropositive/negative defined by 1. MSD-anti-spike titre, 2. MSD-anti-Spike or prior PHE seropositive or total IgG/A/M anti-spike ELISA. 3. PHE sKIDS prior positive (June-July 2020). N.d. – not detected.

**Extended Data Table 2.** Immune characteristics of ELISpot Responding Seronegative donors. **Bold** values indicate values above cut-off. ***Bold-italicised*** indicates values +50% higher than geometric mean.

|  | ELISpot (sfc/10<sup>6</sup> PBMC) |  |  | MSD Serology – AU/ml |  |  |  |  |  |  |  |  |  |  |
| --- | --- | --- | --- | --- | --- | --- | --- | --- | --- | --- | --- | --- | --- | --- |
| Adult /Child | Spike | N/M | Flu | COV-2-Spike | COV-2-RBD | COV-2-NTD | COV2-Nucleocapsid | OC43 Spike | HKU-1 Spike | NL63 Spike | 229e Spike | sKIDS round 1 | sKIDS round 2 | IgG,A,M ELISA |
| Child | **110** | 4 | 172 | 5 | 285 | 2 | 50 | 28273 | **12195** | 2925 | **87935** | Negative | Negative | Negative |
| Child | **243** | 0 | 0 | 12 | 10 | 1 | 546 | 20067 | 1650 | 2012 | 432 | Negative | Negative | Negative |
| Child | **418** | 0 | 766 | 39 | 14 | 0 | 29 | **31263** | **16729** | 3684 | **18035** | Negative |  | Negative |
| Child | **70** | 0 | 796 | 73 | 225 | 2 | **603** | 7360 | 5213 | 1263 | **60299** | Negative | Negative | Negative |
| Child | **142** | 0 | 230 | 40 | 25 | 0 | 59 | 23156 | 5741 | 4402 | 8572 | Negative | Negative | Negative |
| Child | **274** | 0 | 276 | 127 | 97 | 4 | 56 | **53420** | 3992 | **7148** | **14452** |  |  | Negative |
| Child | **330** | 6 | 54 | 134 | 171 | 3 | 241 | 12221 | 1481 | 4213 | **37470** | Negative | Negative | Negative |
| Child | **168** | 0 | 4 | 220 | 47 | 2 | 43 | **29509** | 5580 | 675 | **12144** | Negative | Negative | Negative |
| Child | **742** | 2 | 36 | 25 | 37 | 1 | 93 | 6215 | 2197 | 1425 | 2828 | Negative | Negative | Negative |
| Child | **776** | **32** | 16 | 42 | 40 | 2 | 51 | 2637 | 268 | 305 | 153 | Negative | Negative | Negative |
| Child | **484** | 8 | 136 | 20 | 58 | 1 | 42 | 2610 | 2239 | 1072 | 6684 | Negative |  | Negative |
| Child | **387** | **24** | 104 | 109 | 36 | 1 | **1465** | 9255 | **8513** | 4582 | 5311 | Negative | Negative | Negative |
| Staff | **148** | 0 | 8 | 56 | 323 | 5 | 51 | 43504 | 11602 | 2468 | 28990 | Negative |  | Negative |
| Staff | **62** | 0 | 430 | 69 | 114 | 2 | 46 | **106800** | **26036** | **7039** | **37074** | Negative | Negative | Negative |
| Staff | **496** | 2 | 1246 | 64 | 132 | 0 | 299 | 30002 | 9771 | 2877 | 19008 | Negative | Negative | Negative |
| Staff | **37** | 1 | 19 | 84 | 75 | 3 | 105 | 43170 | 3686 | 4180 | 12688 | Negative | Negative | Negative |
| Staff | **160** | 0 | 392 | 225 | 136 | 7 | **804** | 23610 | 5171 | 826 | 29936 | Negative | Negative | Negative |
| Staff | **1124** | 16 | 374 |  |  |  |  |  |  |  |  | Negative | Negative | Negative |
| Staff | **524** | 2 | 320 | 56 | 39 | 1 | 150 | **62856** | **25802** | 2758 | **33779** | Negative | Negative | Negative |
| Staff | **84** | 4 |  | 65 | 55 | 2 | 278 | **63770** | 9017 | 3795 | 24533 | Negative |  | Negative |
| Staff | 6 | **32** | 96 | 62 | 64 | 12 | 60 | 30596 | 6158 | **5140** | 2549 | Negative | Negative | Negative |
| Staff | 8 | **452** | 64 | 33 | 395 | 8 | 22 | 25250 | 1613 | 692 | **46524** | Negative | Negative | Negative |
| Staff | **428** | 2 |  | 46 | 151 | 1 | 49 | 29970 | **18828** | **4699** | **35577** | Negative |  | Negative |

**Extended Data Table 3.** Antibody reactivity against SARS-CoV-2 spike and RBD from variants of concern in 6+-month convalescent children and adults. *Relative percentage of binding to Wuhan viral protein. †seropositivity defined based on a cut-off of 600 AU/ml.

|  |  | Wuhan |  | B.1.1.7 |  | B1.351 |  | P.1 |  |
| --- | --- | --- | --- | --- | --- | --- | --- | --- | --- |
|  |  | Children | Adults | Children | Adults | Children | Adults | Children | Adults |
| SARS-CoV-2 Spike | Geo. Mean Titre ±SD (AU/ml) | 13138 ±2.1 | 6701 ±2.1 | 11067 ±2.0 | 6351 ±2.1 | 6862 ±2.1 | 3766 ±2.3 | 8477 ±2.1 | 4115 ±2.0 |
|  | Normalised Titre* Median (95%CI) | - | - | 82% (75-89) | 94% (82-109) | 49% (46-59) | 53% (46-73) | 63% (59-66) | 64% (53-74) |
| SARS-CoV-2 RBD | Geo. Mean Titre ±SD (AU/ml) | 4198 ±2.4 | 1954 ±2.4 | 2938 ±2.4 | 1370 ±2.6 | 1299 ±2.3 | 473.3 ±2.4 | 2018 ±2.5 | 691 ±2.5 |
|  | Normalised Titre* Median (95%CI) | - | - | 71% (68-72) | 72% (66-74) | 31% (29-34) | 27% (21-30) | 47% (43-52) | 39% (31-44) |
|  | Seropositive donors<sup>†</sup> (%) | 19/19 (100%) | 17/18 (94%) | 19/19 (100%) | 16/18 (89%) | 16/19 (84%) | 5/18 (28%) | 16/19 (84%) | 8/18 (44%) |

**Extended Method Table 1.**
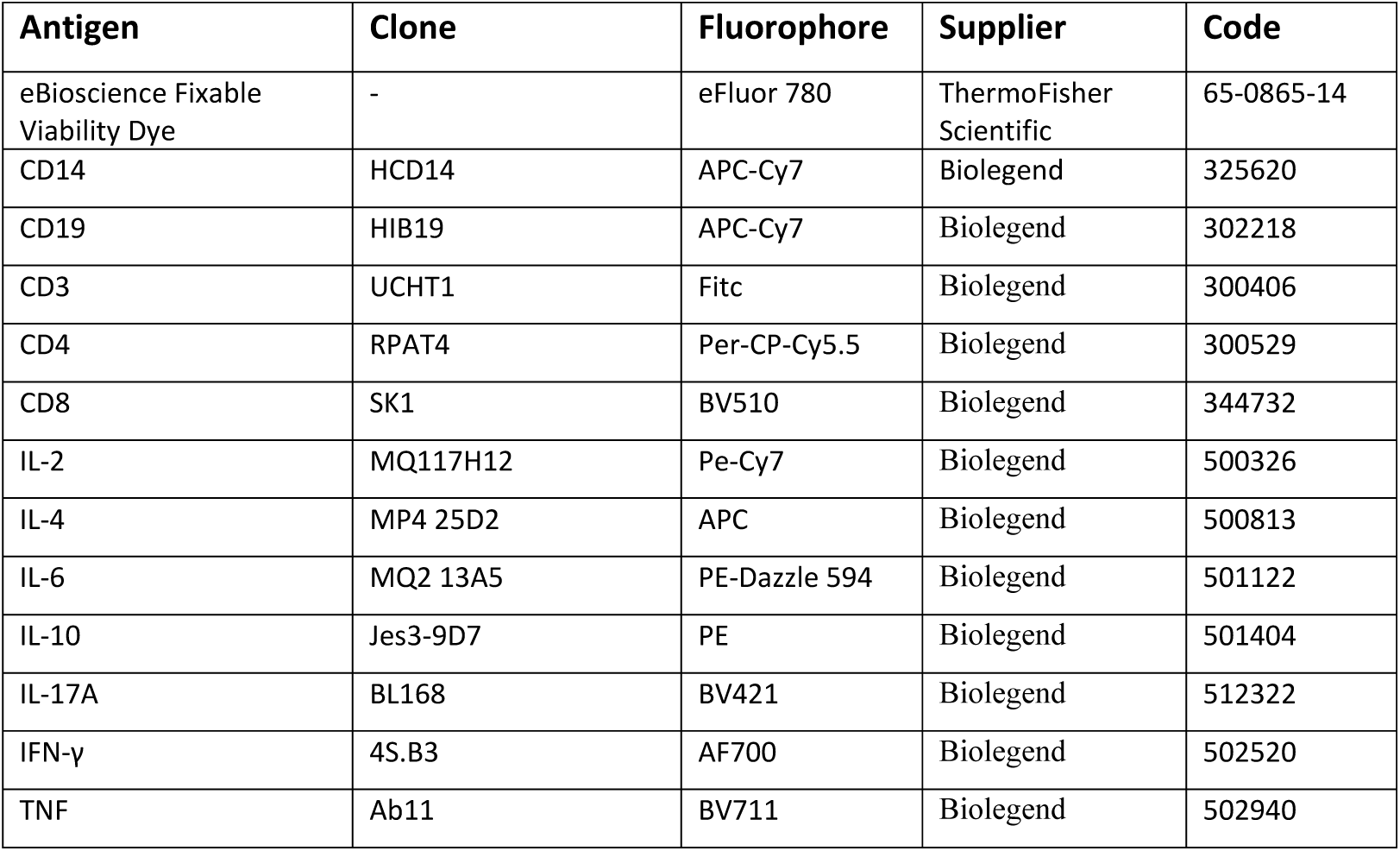
Flow cytometry antibodies.

